# Biomarkers for Atherosclerotic Cardiovascular Events in Rheumatoid Arthritis: Towards Validation of a Biomarker-Enhanced Risk Model

**DOI:** 10.64898/2026.02.18.26346592

**Authors:** Daniel H. Solomon, Leah Santacroce, Jon T. Giles, Pamela Rist, Brendan M. Everett, Katherine P. Liao, Misti Paudel, Nancy A. Shadick, Michael Weinblatt, Joan M. Bathon, Olga Demler

## Abstract

**Background:** Cardiovascular (CV) disease risk is increased in rheumatoid arthritis (RA) and is the leading cause of mortality. Improved CV risk stratification tools in RA could enhance use of preventative care and improve outcomes.

**Methods:** We previously studied biomarkers of CV disease – adiponectin, hsCRP, Lp(a), osteoprotegerin (OPG), high-sensitivity cardiac troponin T (hsTnT), serum amyloid A (SAA), YKL-40, soluble TNF receptor1 (sTNFR1) -- that were associated with CV risk. In the current study, these biomarkers were tested in an unrelated external cohort of RA patients followed at a single academic medical center without a history of CV events. CV events were identified through Medicare and Medicaid administrative data or through medical record review of self-reported events. Biomarkers were assessed at cohort entry among a nested cohort of cases and controls, matched 1:1 on sex and age. Analyses were conducted using conditional logistic regression. We examined whether the candidate biomarkers added to clinical CV risk factors improved model prediction, using the area under the curve (AUC) as well as the net reclassification index (NRI).

**Results:** From a cohort of 1,345 eligible patients with RA, we identified 123 patients with confirmed CV events. Cases and matched controls were typical of RA: median age 63 years, 77% women, RA disease duration 11 years, 72% seropositive, 85% used a biologic or conventional disease modifying anti-rheumatic drug, 58% non-steroidal anti-inflammatory drugs, and 30% oral glucocorticoids. From the candidate biomarkers, LASSO regression selected hsTnT and sTNFR1 as associated with CV events. The AUC for models that included only clinical risk factors was 0.758 (95% CI 0.689-0.829); after adding hsTnT and sTNFR1, the AUC increased to 0.802 (95% CI 0.718-0.998). The NRI of the model with biomarkers was 16.3%, with improvement only observed in patients who did not have CV events during follow-up.

**Conclusions:** Adding selected biomarkers to clinical risk factors enhances the discrimination of models predicting CV events among patients with RA. These risk models require prospective testing to see if they have value in clinical practice decision-making regarding preventative care.

## INTRODUCTION

Cardiovascular disease (CVD) risk stratification has long been recognized as critical to preventing the morbidity and mortality associated with CVD ^1-3^. Although risk stratification for CVD has improved over the last several decades, it has been recognized that general population risk scores do not work well in specific populations, such as chronic kidney disease and rheumatoid arthritis (RA) ^4-6^. These populations experience chronic systemic inflammation, likely contributing to an increased CVD risk and possibly to worse performance of general population risk stratification tools.

In RA, multiple studies demonstrate that risk stratification tools designed for the general population underestimate CVD risk. A study from the Mayo Clinic of 525 patients with RA followed for a mean of 8.4 years illustrated that the Framingham Risk Score (FRS)^7^ consistently underestimated CVD risk in high-risk deciles^4^. Another study found that the regression models with FRS and QRISK2 risk scores had a better fit to data from a non-RA cohort than an RA cohort^8^. Since RA is recognized as an independent risk factor for CVD, several of the risk stratification tools (e.g., QRisk3 and American College of Cardiology (ACC)/American Heart Association (AHA)) consider RA in their list of special populations with enhanced CVD risk^9, 10^. Moreover, the European rheumatology society (EULAR) suggests using the SCORE risk tool and multiplying by 1.5 for some groups with RA^11^. While these efforts acknowledge the problems with existing CVD risk stratification tools, they do not use an evidence-based approach to derive and/or validate a better tool for individuals with RA.

There have been several prior efforts to derive and validate CVD risk stratification tools specifically for RA. One set of studies derived a risk tool starting with traditional risk factors found in the FRS and then tested whether adding in RA-specific factors improved model performance^12^. The original derivation study used a large US-based RA registry and found small improvements in the c-statistic and a net reclassification index (NRI) of 40% (95% CI 37-44%) by adding four RA clinical variables: glucocorticoid use, disease duration ≥ 10 years, moderate disease activity, and functional disability. This risk tool has been further validated in several external cohorts^13, 14^. However, this tool did not attempt to include biomarkers of CVD. Another prior effort to assess CVD risk among Medicare patients with RA used a proprietary weighting of biomarkers (from the Multi-Biomarker Disease Activity, MBDA panel) plus age, diabetes, hypertension, smoking, CVD history, leptin, MMP3, and sTNFR1 as variables in a regression model^15^. They found that this group of variables was associated with future CVD events over a 3-year follow-up. As well, it seemed to replicate in an external cohort^16^. However, the weights used in the MBDA are not described, the 3-year time horizon is very brief, and the C-index was relatively low (0.72).

In the current study, we examined several known CVD biomarkers that have also been found to be associated with RA. These biomarkers were studied in a prior RA cohort and found to be associated with FDG PET/CT vascular inflammation (measured by target-to-background ratio). Thus, this study aimed to assess their relationship with CVD events assessed in a large independent RA cohort and to determine if adding these biomarkers to a risk prediction algorithm would improve model performance.

## METHODS

### Study Population and Design

We designed a nested case-control study with atherosclerotic CVD events as the case-defining outcome. The study was conducted in the context of the Brigham and Women’s Hospital (BWH) Rheumatoid Arthritis Sequential Study (BRASS), which is a single-center longitudinal cohort that has been previously described^17^.

Participants in BRASS all had a clinical diagnosis of RA from a board-certified rheumatologist. Follow-up visits occurred at least twice each year, with visits including RA disease activity assessments, comorbidity and medication inventories, and biospecimen collections.

We identified cases of incident CV events from the BRASS cohort (see below). Controls were matched to cases based on age (+/-5 years) and sex at BRASS cohort entry. All BRASS participants without known CVD at baseline, between ages 35 and 90, were eligible to be cases or controls. Incidence density sampling of cases and controls was used such that a participant without a known CVD event could be selected as a control up until the time they became a case.

### Cardiovascular Outcomes: Case Definition

The BRASS cohort was not designed to assess CVD, but participants were asked about the presence of coronary artery disease, strokes, and peripheral vascular disease at baseline; as well, they were asked about any atherosclerotic CVD events (myocardial infarction, coronary re-vascularization, unstable angina, stroke, or transient ischemic attack) at least annually. Many, but not all, BRASS participants receive most of their care at BWH or an affiliated hospital, allowing us to review medical records. Furthermore, administrative insurance data (such as diagnosis and procedure codes) are available for BRASS participants who have their health care paid by Medicare and/or Medicaid. We used self-report, medical records, and administrative data to define the composite outcome of ASCVD event.

The outcome for these analyses required medical record evidence or administrative data evidence for a non-fatal myocardial infarction (MI), non-fatal stroke or transient ischemic attack (TIA), unstable angina requiring hospitalization, or re-vascularization (percutaneous coronary intervention or coronary artery bypass graft), or CV death. This composite is similar to 4-point MACE^18^. The administrative data definitions of these components all have high positive and negative predictive values (> 80%)^19-21^. The medical record review was conducted by an experienced CVD outcome adjudicator (BME), using standard definitions.

### Biomarker Assessment

We focused on testing biomarkers that could be clinically useful and relatively easily tested in a clinical laboratory. In prior work, we tested 25 biomarkers for their association with atherosclerotic CVD risk, assessed using a FDG PET/CT test of vascular inflammation^22^. The target-to-background ratio (TBR) was estimated using readings from an experienced reading core that assessed bilateral carotid and aorta images, using standard methods^23^. The prior work identified six biomarkers associated with TBR: adiponectin, high sensitivity C-reactive protein (hsCRP), osteoprotegerin (OPG), serum amyloid A (SAA), soluble TNF receptor 1 (sTNFR1), and YKL-40^22^. We added high sensitivity troponin T (hsTnT) and lipoprotein a (Lp(a)) to the list of potential biomarkers, as there are strong data to support their utility as CV biomarkers outside of our prior study^24, 25^.

These eight biomarkers were tested for their ability to predict future CVD events in BRASS study participants without known CVD. Biospecimens had been collected at entry into BRASS or at the one-year visit and were stored at −80°C. They were retrieved and sent to a CLIA-certified clinical laboratory (Boston Children’s Hospital, Boston MA). Biospecimens from cases and controls were assayed in random order and the laboratory personnel were blinded to case status.

Biomarkers were run using commercially available ELISA kits (Roche diagnostics). The laboratory used commercial quality control (QC) material where available to obtain different levels of each biomarker; they also used their own pool of frozen serum/plasma for QC. The BRASS samples were only run after all QC specimen levels were within range. Then, after every 50 samples, QC was repeated. When running ELISAs, the samples were run in 96-well plates. Each plate can run at most 38 samples in duplicate, besides the levels of QC and the standard curve. Every plate had its own set of controls, and the samples were run in duplicate and averaged to monitor any pipetting issues.

### Other Cardiovascular Risk Factors

Since the goal of this work is to assess whether biomarkers improve CV event risk stratification among patients with RA, we considered traditional CVD risk factors: age, sex, diabetes, hypertension, hyperlipidemia, current tobacco use, and BMI. Actual lipid parameters were not available on most patients, making imputation impossible and prohibiting use of standard CV risk scores, such as PREVENT^26^ or the AHA/ACC pooled cohort equation^27^. We used two alternative approaches with traditional CVD risk factors. First, we used traditional CVD risk factors, including variables listed in Table 1. RA clinical variables, selected based on prior work^12^, were added to these traditional CVD risk factors. Second, we used a previously developed CVD risk score modified for RA, the “Extended Risk Score for RA” (ERS-RA)^12^. The ERS-RA contains traditional CVD risk factors plus four RA clinical variables: RA disease duration ≥ 10 years (yes/no), use of oral glucocorticoids (yes/no), clinical disease activity index of moderate or high disease (yes/no)^28^, and a modified health assessment questionnaire > 0.5 (yes/no)^29^. In the original work deriving the ERS-RA, these four variables were among 15 RA clinical variables tested and were found to enhance the C-index. As noted above, the ERS-RA has been externally validated in other RA cohorts^13, 14^.

**Table 1:** Baseline Characteristics of Cases and Controls in the Rheumatoid Arthritis Study Population.

|  | Total* | Controls | Cases* |
| --- | --- | --- | --- |
|  | N = 246 | N = 123 | N = 123 |
|  | *Median (25%, 75%) or n (%)* |  |  |
| Age, years | 63 (58, 69) | 63 (56, 68) | 63 (59, 70) |
| Sex, female | 190 (77%) | 95 (77%) | 95 (77%) |
| **Cardiovascular factors** |  |  |  |
| Hypertension | 115 (47%) | 54 (44%) | 61 (50%) |
| Hyperlipidemia | 83 (34%) | 40 (33%) | 43 (35%) |
| Diabetes | 24 (9.8%) | 8 (6.5%) | 16 (13%) |
| Tobacco use, current | 16 (6.5%) | 3 (2.4%) | 13 (11%) |
| Statin use | 51 (21%) | 25 (20%) | 26 (21%) |
| Anti-hypertensive use | 106 (43%) | 48 (39%) | 58 (47%) |
| Systolic blood pressure, mmHg | 135 (120, 142) | 131 (120, 140) | 137 (122, 147) |
| Body mass index, kg/m<sup>2</sup> | 26.4 (23.5, 29.9) | 26.1 (23.5, 29.2) | 27.1 (23.3, 30.5) |
| **Rheumatoid arthritis factors** |  |  |  |
| NSAID use | 143 (58%) | 67 (54%) | 76 (62%) |
| Oral glucocorticoid use | 73 (30%) | 28 (23%) | 45 (37%) |
| DMARD use, any | 210 (85%) | 110 (89%) | 100 (81%) |
| RA clinical disease activity index | 20 (11, 36) | 22 (12, 36) | 18 (10, 34) |
| mHAQ-DI | 0.6 (0.2, 0.9) | 0.5 (0.2, 0.8) | 0.7 (0.3, 1.1) |
| Disease duration, years | 11 (4, 21) | 11 (4, 19) | 13 (5, 23) |
| Seropositive | 180 (73%) | 82 (67%) | 98 (80%) |

Abbreviations: RA, rheumatoid arthritis; NSAID, nonsteroidal anti-inflammatory drugs; DMARD, disease-modifying anti-rheumatic drugs, including biologic and non-biologic; mHAQ-DI, modified health assessment questionnaire-disability index. Cardiovascular and RA factors are defined as present or absent. Seropositive was defined based on either a rheumatoid factor or anti-CCP antibody that was considered positive. \*Cases are defined using the atherosclerotic CV composite endpoint definition described in the text.

### Statistical Analyses

After defining cases, these patients were matched 1:1 with controls. Baseline characteristics were compared between cases and controls using the median (interquartile range, IQR) for continuous variables and relevant categories of non-continuous variables. The distributions of each biomarker were examined, values above or below the limit of detection were imputed, outliers were scaled, and values were standardized using Z-scores to reduce scale-related differences and enable direct comparison.

Conditional logistic regression models were then examined. First, the association of each biomarker with CVD event was tested individually with age forced into all models. Two biomarkers were found to have p< 0.05 in age-adjusted bivariate models and retained for multivariable testing with clinical risk factors. These two biomarkers were then added into the primary analyses: one model with traditional cardiovascular and rheumatoid arthritis risk factors, and another model with the ERS-RA risk score.

Model fit with and without biomarkers was examined using the area under the receiver operating characteristic curve (AUC). The AUC was estimated using a semi-parametric method that accounts for the matched case-control design that uses a semi-parametric approach to estimate AUC for case-control studies. 95% confidence intervals (CIs) around the AUC were estimated with bootstrapping with 100 runs. The Akaike Information Criterion (AIC; smaller is better) was estimated, and the likelihood ratio test was used to compare the fit of different models. Finally, we estimated the net reclassification indices (NRI) for the models with biomarkers versus without. The 10-year CV event risk categories considered in estimating NRI were ≤ 5%, >5% to ≤ 7.5%, >7.5% - ≤ 20%, and >20%. The NRI was estimated overall and for cases and controls separately.

All analyses were conducted using R Version 4.5.0

## RESULTS

The study population was drawn from 1,581 patients with RA in the BRASS registry. After eliminating those who reported a CVD event prior to enrollment (n = 213) and had a self-reported CVD event that could not be confirmed during follow up (n = 80), we were left with a cohort of 1,288 patients which we call the parent cohort. The median follow-up for the parent cohort was 5.03 (IQR 2.95 – 9.99) years. We identified 123 patients from the parent cohort with CVD events who had stored baseline blood samples available for biomarker measurement. They were matched based on sex and age to an equal number of controls. The baseline characteristics of cases and matched controls are shown in **Table 1** and are typical for RA patients: median age 63 years, 77% women, RA disease duration 11 years, 72% seropositive, 85% used a biologic or conventional disease-modifying anti-rheumatic drug, 58% non-steroidal anti-inflammatory drugs, and 30% oral glucocorticoids. CV risk factors were consistently more common in cases versus controls. The incidence rate for CVD events in the parent cohort was 13.55 (95% CI 11.35 – 16.17) per 1,000 person years. The distribution of time until events is shown in **Supplemental Figure 1**, with most events (67%) occurring within 10 years of entry into the cohort.

The median biomarker values for cases and controls are shown in **Figure 1** and **Supplemental Table 1**. The standardized values (Z-scores) of the biomarkers were tested in matched conditional logistic regression models, one at a time; age was forced into all models (see **Table 2**). hsTnT and sTNFR1 were significantly associated with CVD events in the age-adjusted models and were retained for testing in multivariable regression models.

**Figure 1.**
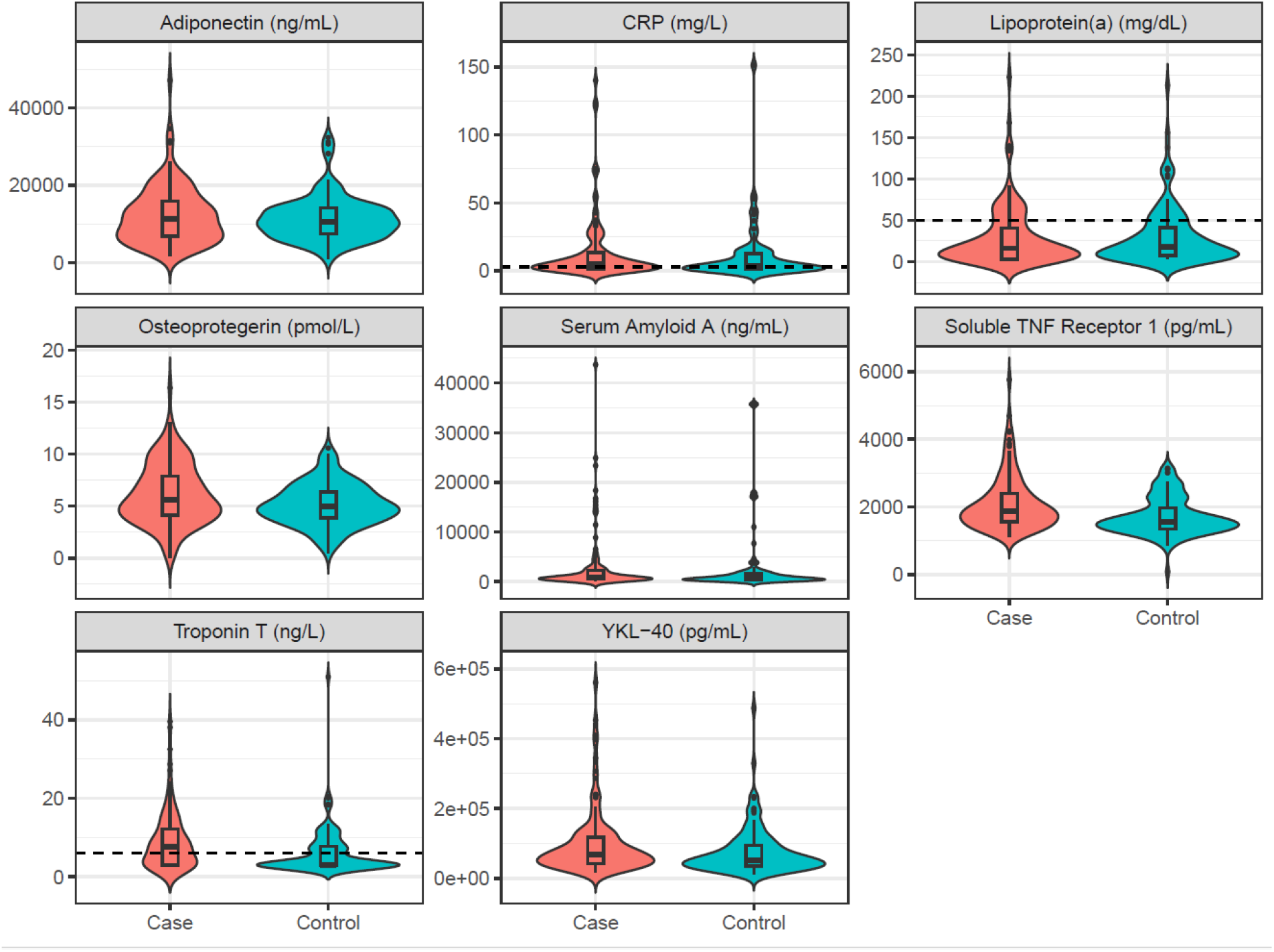
The distributions of biomarker values are shown in these violin plots with the median and interquartile ranges noted. The dashed lines on hsCRP, lipoprotein(a) and troponin T denote thresholds for elevated values. These include the following: hsCRP > 3mg/L; lipoprotein(a) ≥ 50mg/dL; and troponin T > 6ng/L. The medians and interquartile ranges for these biomarkers are shown in **Supplemental Table 1**.

**Table 2:**
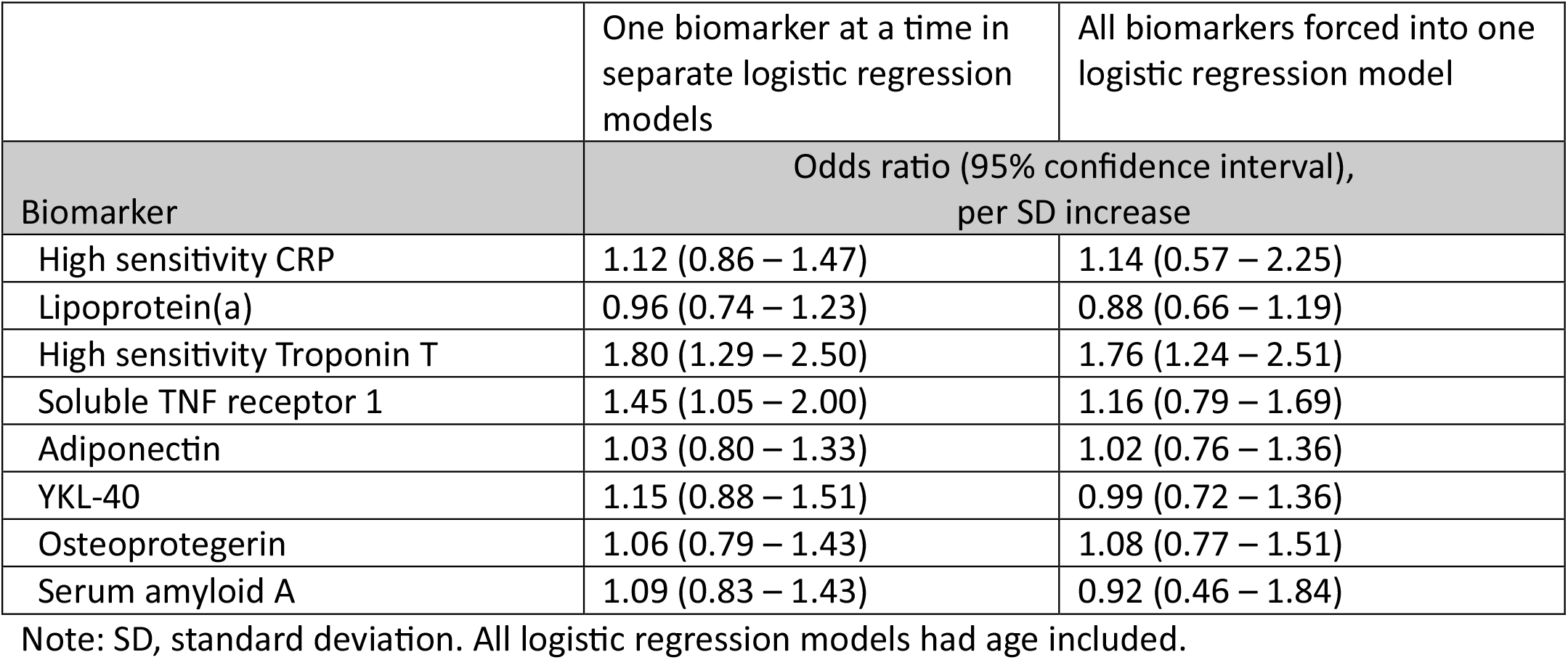
Regression Analyses Testing Association Between Individual Biomarkers and Atherosclerotic Cardiovascular Events.

Using conditional logistic regression models, we tested the incremental model fit, including only clinical factors and then adding the two retained biomarkers: hsTnT and sTNFR1 (see **Table 3**). The AUC improved when the two biomarkers were added to the clinical risk factor model: AUC 0.758 (95% CI 0.689-0.829) for clinical risk factors versus AUC 0.802 (95% CI 0.718-0.998) for the model also including the two biomarkers (likelihood ratio test p-value 0.004). The AIC also demonstrated significant improvement: 320.5 versus 313.3 for model with biomarkers. The improvements for the ERS-RA model were less obvious: AUC 0.743 (95% CI 0.682-0.813) versus AUC 0.756 (95% CI0.706-0.992) for the model with biomarkers (likelihood ratio test p-value 0.002; AIC 143.49 versus 136.39 with biomarkers.

**Table 3:**
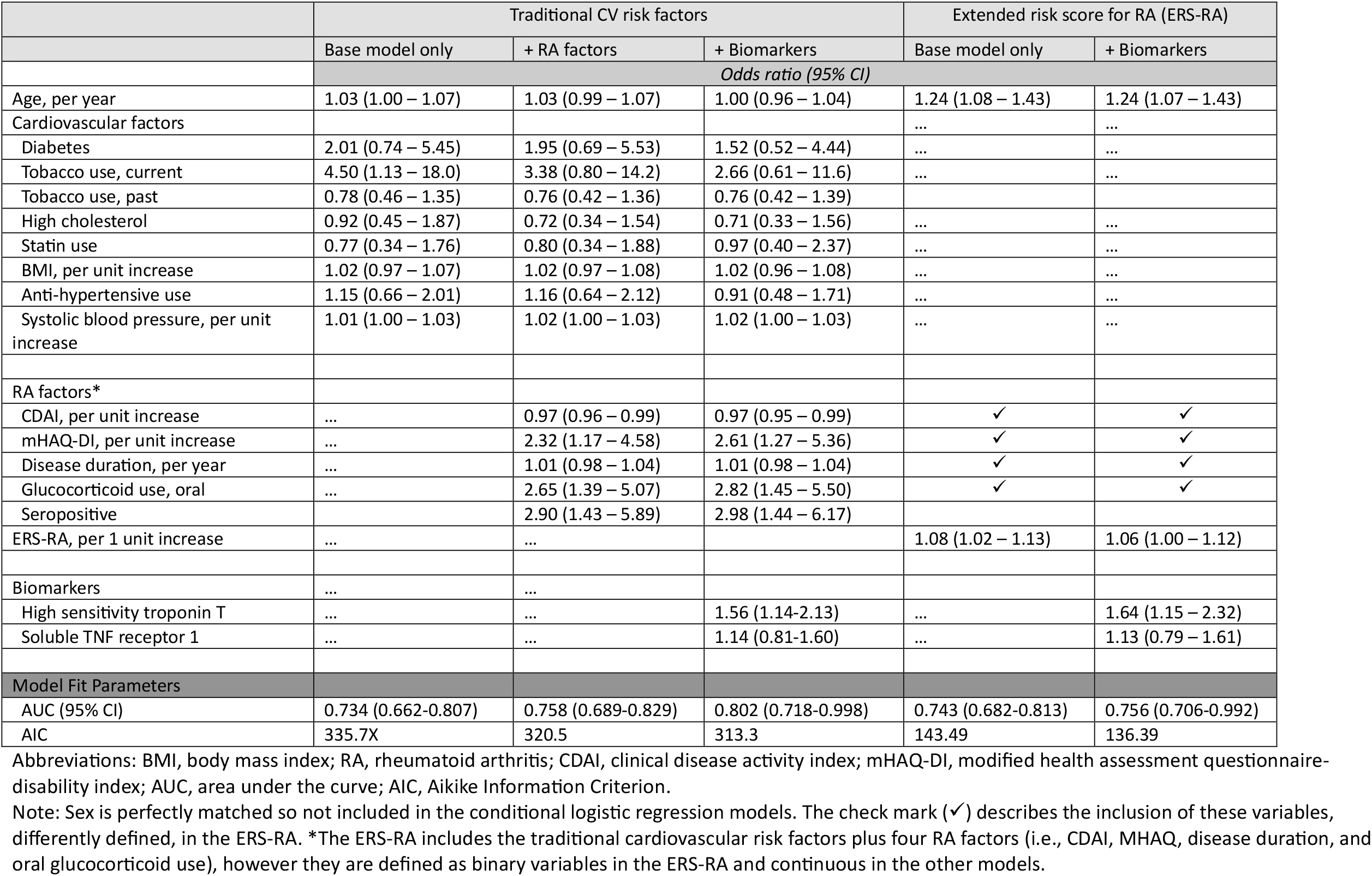
Conditional Logistic Regression Analyses Testing Association Between Clinical Risk Factors, Biomarkers, and Atherosclerotic Cardiovascular Events.

To better assess whether the model improvements with biomarkers resulted in better clinical classification of risk, we assessed the NRI (see **Supplemental Table 2**). The NRI for the clinical risk factor model (top of **Table**) showed an NRI = 16.3% (95% CI 1.0-31.8) when including biomarkers, with all of the improvement in reclassification among the controls, and no improvement was seen for cases (see **Supplemental Table 3a**). While the NRI for the ERS-RA was slightly larger (NRI = 17.9%, 95% CI 0.8-34.7%), the reclassification was negative (worse) for cases (−11.4%) but strongly positive for controls (29.3%) (see **Supplemental Table 3b**).

## DISCUSSION

Previous work has observed the inaccuracy of general population CV risk models in specific high-risk groups, such as patients with RA^4^. Past efforts have attempted to improve the accuracy of general population risk models by using a multiplier^30^, adding in RA clinical variables^12^, or using a proprietary multi-biomarker panel^16^. The prior work all had important limitations. Thus, we tested the value of candidate biomarkers in predicting CV risk among patients with RA. This was done in the setting of a large RA registry with validated atherosclerotic CV events. We found that two biomarkers – hsTnT and sTNFR1 -- significantly improved model fit compared with models containing only clinical variables. The NRI demonstrated improvements that have potential clinical importance for targeting preventative CV care, if further replicated.

Model fit was strong after adding in biomarkers, as demonstrated by improvements in the AUC, AIC and NRI. This suggests the potential for these models to be used in clinical practice if results are confirmed in additional external cohorts. Such studies should assess not only model fit, but whether the biomarker-enhanced models help categorize patients into risk groups requiring or not requiring clinical interventions, such as further testing or CV risk factor management. In other words, does the use of biomarkers more accurately identify patients with RA who should or should not have coronary artery calcium (CAC) scanning for better risk stratification? Alternatively, biomarkers might help categorize CV risk to identify who should receive aggressive preventative treatments.

How might one consider further testing these biomarkers for CVD risk among patients with RA? One possibility is to identify patients who appear at low or high CV event risk by accepted clinical risk scores (i.e., using PREVENT^31^ or the pooled cohort equation, PCE^32^). These patients could then have biomarkers tested and undergo a CAC scan. Using the CAC score as the gold standard, established risk models (e.g., PREVENT or PCE) could be compared with models that also include biomarkers. The key question to answer is whether models containing biomarkers better predict CAC scores that would or would not prompt preventative treatment.

Prior literature supports the biologic plausibility of hsTnT and sTNFR1 being true biomarkers of atherosclerotic CVD in RA. hsTnT is a protein released from cardiac myocytes during times of acute injury, but it can also be found at low levels in the blood by assays during subclinical cardiac injury^33^. Prior studies demonstrate that hsTnT is elevated in microvascular disease, such as RA^34-36^. In the ARIC study, persons with hsTnT levels in the highest quintile had an approximately three-fold increased risk of a future CV event^37^. sTNFR1 is highly correlated with TNF activation and is stable in circulation. In addition to being a good marker of chronic inflammation, sTNFR1 was strongly associated with future CV events in a general population cohort with nearly 4,000 patients^38^, with hazard ratios per standard deviation increase in sTNFR1 between 1.1 and 1.3 in derivation and validation cohorts. Prior studies of these two biomarkers in patients with RA clearly demonstrate increased CV event risk^24^ and all-cause mortality^39^. Data from the current study, combined with previous literature, strongly support the potential for incorporating these biomarkers into CV risk models in RA.

The limitations of the current study are important to recognize. First, relatively few atherosclerotic CV events were included in this study, limiting the statistical power of the regression models. Second, the external validation cohort patients all came from one rheumatology practice, limiting generalizability. Third, a limited number of biomarkers were tested based on their performance in models with vascular inflammation (FDG PET/CT) as the endpoint; however, they had all been tested in prior work^22^, so the a priori probability of their significance was strong. Finally, there was no direct measure of the lipid profile. Strengths of the study need to be highlighted as well. The regression models used adjudicated CV events, rather than a surrogate or patient-reported outcome. All data were collected prospectively with long follow-up. Finally, the biospecimens were collected at timepoints before the CV events and measured using reproducible commercial assays.

In conclusion, we have tested several biomarkers to determine if they improve model fit statistics in regression analyses with CV events as the outcome among a cohort of patients with RA. From a group of previously tested biomarkers, we found two – hsTnT and sTNFR1 – improved the model fit compared to a model with known clinical CV risk factors and RA clinical variables previously shown to improve CV risk prediction. While these results are encouraging, further testing of these biomarkers in additional RA cohorts to determine if they enhance risk prediction over the PREVENT risk score would be useful. As well, testing them using a CAC score as the outcome would allow us to determine if they help with CV prevention decision-making.

## Data Availability

Data can be made available to researchers with an approved IRB protocol

